# The Effects of Stress on Academic Performance Among High School Students in Lusaka

**DOI:** 10.1101/2021.05.26.21257875

**Authors:** Miyoba Hachintu, Friday Kasisi

## Abstract

**OBJECTIVE:** This study aimed to identify the various factors associated with stress and how those factors affect academic performance among high school students in Lusaka

**BACKGROUND:** The word stress brings about the thoughts of depression, anxiety and other potentially life-threatening conditions in the mind of an individual. Every person is exposed to stress at some point in their lives, and it is an inevitable part of a student’s life as it takes a toll on their emotional well-being, academic performance as well as their physical health. Different factors cause stress among students. These include relationships with family and friends, examinations and deadlines, poor time management, social media, financial instability, future career thoughts, depression, anxiety and many more. Educational environments are extremely competitive, and students must be able to deal with academic related stress by relying on their coping capabilities. Most students are unable to cope with stress. This leads to different behavioural patterns such as alcoholism and substance abuse such as “dagga” (marijuana) and codeine in order to escape the harsh reality. This eventually leads to absentia in school and an increased number of school drop outs. Stress is known to be the source of many problems among youth and its effects can be as toxic as suicide. Therefore, it’s important that parents or guardians, teachers, the students themselves and the entire nation unite and ensure that supportive data is communicated to students for them to cope with their stress levels in a responsible manner.

## METHODS

A cross-sectional research design was employed in this study. A total of 232 students were selected from a population of 2, 322. A probability proportionate stratified sampling technique was used in this study. The first stage involves stratification of the population based on gender. The aim is to attain a representative sample of each stratum (Saunders et al., 2012). The second stage employs the proportionate sampling technique to determine the actual sample to be drawn from each group which is a fraction of 1/10. The third stage employs random sampling of students that will participate in the study using a list of registered students from the three schools as the sampling frame. 40 from 400 at Lake Road PTA School, 102 from 1,022 at Kabulonga Girls Secondary School and 90 from 900 at Kabulonga Boys Secondary School.

A semi-structured self-administered questionnaire was used to collect primary data for the study. The questionnaire was in form of a Student Academic Stress Scale (SASS) and was divided into five main parts. The first part included the demographics of the students. The second dimension, participants responded to each question concerning the causes of stress by ticking a suitable answer on the Likert Scale with five measurement: Most Stressful (1); Stressful (2); Moderately Stressful (3); Less Stressful (4); and Not Stressful at all (5). The third dimension consisted of questions that were aimed at identifying how stress affected academic performance of students. The fourth dimension contained a few open-ended questions asking students how best they thought they could be helped when they were overwhelmed with stress. The last part consisted of various ways that respondents coped with stress which required them to tick suitable answers.

Secondary data was also obtained through various international journals and articles on the topic. Academic performance was measured using student report forms which were provided by the head teachers of the students. Descriptive statistics such as frequencies and percentages were used to summarise demographic information of the respondents. Inferential statistics such as chi-square tests with a significant level of 5% was used to test the association between variables.

## RESULTS

The analysis of this research was purely quantitative. It was done by calculating the frequency, percentage and the p-values of the factors of stress among students, how stress affects academic performance.

**Table 1.** Association between factors of stress and academic performance

| Variables | Performance |  | P-Value |
| --- | --- | --- | --- |
|  | Passed | Failed |  |
| Too Many Assignments |  |  |  |
| Yes | 51 (36.7%) | 67 (72.0%) | p <0.001 |
| No | 88 (63.3%) | 26 (28.0%) |  |
| **Student Competition** |  |  |  |
| Yes | 63 (45.3%) | 28 (30.1%) | p=0.020 |
| No | 76 (54.7%) | 65 (69.9%) |  |
| **Content of Subjects** |  |  |  |
| Yes | 86 (61.9%) | 73 (78.5%) | p=0.008 |
| No | 53 (38.1%) | 20 (21.5%) |  |
| **Teaching Style of teachers** |  |  |  |
| Yes | 96 (69.1%) | 60 (64.5%) | p=0.469 |
| No | 43 (30.9%) | 33 (35.5%) |  |
| **Poor Relationship with teachers** |  |  |  |
| Yes | 89 (64.0%) | 76 (81.7%) | p=0.004 |
| No | 50 (36.0%) | 17(18.3%) |  |
| **Absence of teachers** |  |  |  |
| Yes | 92 (66.2%) | 66 (71.0) | p=0.444 |
| No | 47 (33.8) | 27 (29.0) |  |
| **Failing a major subject** |  |  |  |
| Yes | 77 (55.4%) | 67 (72.0%) | p=0.013 |
| No | 62 (44.6%) | 25 (26.9%) |  |
| **Family Relationships** |  |  |  |
| Yes | 75 (54.0%) | 48 (51.6%) | p=0.726 |
| No | 64 (46.0%) | 45 (48.4%) |  |
| **Learning Environment** |  |  |  |
| Yes | 83 (59.7%) | 57 (61.3%) | p=0.810 |
| No | 56 (40.3%) | 36 (38.7%) |  |
| **Domestic Responsibility** |  |  |  |
| Yes | 100 (71.9%) | 68 (73.1%) | p=0.844 |
| No | 39 (28.1%) | 25 (26.9%) |  |
| **Physical Health** |  |  |  |
| Yes | 66 (47.5%) | 46 (49.5%) | p=0.767 |
| No | 73 (52.5%) | 47 (50.5%) |  |
| **Examinations** |  |  |  |
| Yes | 106 (76.3%) | 83(89.2%) | p=0.013 |
| No | 33(23.7%) | 10(10.8%) |  |
| **Lack of Quality Sleep** |  |  |  |
| Yes | 119 (85.6%) | 67(72.0%) | p=0.011 |
| No | 20 (14.4%) | 26(28.0%) |  |
| **Limited Time** |  |  |  |
| Yes | 115 (82.7%) | 74 (79.6%) | p=0.543 |
| No | 24 (17.3%) | 19 (20.4%) |  |

The table shows the association of the possible stressors with academic performance of students. The stressors that have a positive relationship with academic performance and gave a significant result are too many assignments, student competition, content of subjects, poor relationship with teachers, failing a major subject, examinations and lack of quality sleep.

## CONCLUSION

This narrative review highlights that stress is a major concern for secondary school students. This study identifies various factors of stress among students being academic workload, student competition, difficulty understanding the content of subjects, poor relationships with teachers, lack of quality sleep, limited time to adequately prepare schoolwork and examinations. It also identifies how stress affects academic performance through negative attitudes towards school, strained relationships with teachers, failed subjects and lack of confidence in their academic work. These in turn lead students to engage in different strategies or modes of coping with the stress such as problem-focused or emotion-focused strategies as stated by Lazarus and Folkman (1984).

Findings show that to a large extent, stress affects students negatively. Therefore, it is vital for schools to understand the various sources of stress among their students and this implies putting effective stress overcoming measures in place such as enabling counsellors to create awareness and tailor-made intervention programs that are relevant to students’ academic success and general life.

## LIMITATIONS OF THE STUDY

There was a challenge of collecting data due to COVID 19 restrictions in the schools.

## Data Availability

All data generated or analysed during this study are included in this published article (and its supllemnatry information files).

https://www.researchgate.net

## Notes

### Competing Interest Statement

The authors have declared no competing interest.

### Clinical Trial

The study used a cross-sectional research design.

### Funding Statement

This resarch received no external funding

### Author Declarations

Institutional Review Board Statement: Intuitional authorization was obtained from the University of Lusaka, School of Medicine and Health Sciences Research Ethics Committee (Reference: IORG0010092/MPH19114647). National Health Research Authority (NHRA) granted permission (Ref No: NHRA00005/09/01/2021) in line with the Act of Parliament (No. 2 of 2013) which mandatory requires all researchers to submit their research protocols to the NHRA upon receipt of approval from a Research Ethics Committee or an Institutional Review Board

